# fMRI analysis parameters affect the concordance with TMS in noninvasive speech mapping

**DOI:** 10.64898/2026.01.29.26345106

**Authors:** J Gogulski, S Autti, M Vasileiadi, M Tik, S Vaalto, H Renvall, M Liljeström, P Lioumis

## Abstract

**Background:** Speech cortical mapping (SCM) conducted with widely available functional MRI (fMRI) can yield divergent results compared to the more commonly used navigated TMS (nTMS). The impact of specific fMRI task paradigms and preprocessing choices on reaching similarity with nTMS has not been explored before.

**Objective:** To test how the fMRI experimental task and spatial smoothing of the data compare with nTMS-based results, to subsequently increase the reliability of object naming fMRI for SCM.

**Methods:** Thirteen healthy, right-handed Finnish speakers underwent an nTMS-based SCM experiment in which the left hemisphere was stimulated while the subjects overtly named common visually presented stimuli. Standard as well as magnetoencephalography-informed picture-to-TMS intervals were applied. The same participants completed fMRI with overt naming, silent naming, and observation tasks on the same stimuli, analyzed with 0-, 3-, and 6-mm spatial smoothing. nTMS-based error and non-error sites were converted to volumetric density maps, and error-specific maps were derived by subtracting non-error from error density. Spatial similarity between binarized fMRI maps and nTMS maps was quantified using Jaccard index. Within-session fMRI reliability was estimated with voxel- and subjectwise concordance correlation coefficients across two separate runs conducted on the same day.

**Results:** Similarity between fMRI and nTMS maps was overall low but depended significantly on data smoothing. Within subjects, mean error-specific Jaccard index was 0.036, with most individuals showing maximal similarity at 6 mm of smoothing. The fMRI task resulting in highest similarity with the nTMS map varied across participants, but at the group level, silent naming with 6-mm smoothing yielded the best correspondence. In general, within-session fMRI reliability increased with greater smoothing.

**Conclusion:** The amount of applied fMRI data smoothing shapes the agreement of fMRI and nTMS maps during SCM. Silent naming fMRI combined with 6-mm data smoothing yielded the highest overlap with nTMS maps, yet the effect of the experimental task was statistically non-significant and the absolute similarity of the maps remained low. These results underline the different views to brain functions provided by direct perturbation of neural functions vs. blood-oxygenation based fMRI, and offer practical guidance when combining fMRI with nTMS in noninvasive speech cortical mapping.

**Highlights:**

- Correspondence of fMRI and TMS speech cortical mapping results varied across individuals
- Concordance between the methods was generally low and depended on the fMRI data smoothing
- Silent naming task in fMRI, combined with 6-mm data smoothing, yielded highest similarity to nTMS
- Within-session fMRI reliability increased with greater smoothing

## 1. Introduction

Preoperative speech cortical mapping (SCM) is commonly pursued in patients with tumors or epilepsy whose lesions are close to regions critical for speech production (Leone et al., 2025). Among noninvasive techniques, navigated transcranial magnetic stimulation (nTMS) is frequently applied to the cortical surface during object naming to pin-point language-critical regions eliciting transient stimulation-related naming errors (Lefaucheur & Picht, 2016; Lioumis et al., 2012; Raffa et al., 2019). nTMS mapping correlates well with direct cortical stimulation (DCS), the gold standard for SCM accuracy (Krieg et al., 2017). Interpretability of nTMS-based SCM is, however, affected by several factors. First, optimal stimulation timing relative to stimulus onset likely varies across individuals and linguistic tasks (Hari et al., 2000; Pulvermüller et al., 2003; Tatsuno & Sakai, 2005). Magnetoencephalography (MEG) studies of object naming have identified inter-individual variability in the timing of neural activation and response times (Liljeström et al., 2009, 2015), supporting the idea that individualized latencies are preferable over fixed timing for all participants (Lioumis et al., 2023). Furthermore, the spatial sampling density in TMS mapping is constrained by session duration and operator choices, which can influence the balance of false positive and false negative findings with respect to DCS (Jeltema et al., 2021). To mitigate both timing- and sampling-density-related uncertainties in nTMS-based SCM, it might be useful to leverage additional imaging to inform when and where to stimulate, rather than relying on the TMS operator’s neurophysiological and neuroanatomical knowledge alone.

Functional MRI (fMRI) is a widely available method that can also be used for localization of language-related areas (Branco et al., 2016; Brennan et al., 2016; Ille et al., 2015; Rutten et al., 2002; Szaflarski et al., 2008). It covers the whole brain in a single scanning session and is less operator-dependent than nTMS. On the other hand, single-subject fMRI results can be noisy, and are generally sensitive to the experimental task and analytical choices, e.g., spatial smoothing (Ning et al., 2019), limiting their reliability and clinical utility. Currently, fMRI results are typically limited to assessing language lateralization before neurosurgery. Importantly, fMRI, which measures hemodynamic activation, probes different aspects of language organization than TMS and DCS which cause perturbation of the speech process via neuronal activation: Thus, the mapping results can be expected to differ between methods (Gogulski et al., 2025).

Noninvasive brain imaging methods can provide meaningful complementary information. fMRI may provide evidence for spatial prioritization of the perturbation due to its high spatial resolution, while MEG could potentially inform of subject-specific temporal stimulation windows during naming in addition to spatial information from the superficial parts of the cortex (Ala-Salomäki et al., 2021; Grummich et al., 2006; Liljeström et al., 2009; Tarapore & Nagarajan, 2017). fMRI-guided TMS is feasible in language mapping (Vasileiadi, Schuler, et al., 2023; Vasileiadi et al., 2026). At the same time, fMRI results are of correlational nature, and depend on task choice (Benjamin et al., 2018) and analysis parameters. For example, spatial smoothing affects the intraindividual reproducibility (Ning et al., 2019; Vasileiadi, Woletz, et al., 2023). Prior reports have compared nTMS-based language mapping with fMRI activation (Ille et al., 2015; Schiller et al., 2020a), but to our knowledge, no study has systematically evaluated how fMRI experimental task and size of smoothing kernel influence within-subject concordance with nTMS-derived speech maps.

In this study, we mapped language areas in healthy, right-handed Finnish speakers using individualized nTMS (N=13, one of which did not participate in the fMRI experiment) and task-based fMRI (N=13, one of which did not participate in the nTMS experiment). nTMS mapping targeted classical speech-related areas on the left hemisphere, as well as nearby non-speech-related areas. We applied several standard stimulation latencies ranging between 0-500 ms, as well as stimulation latencies informed by each participant’s MEG object-naming responses. We thus used a wide time window for picture-to-TMS interval (PTI), rather than a single stimulation latency for TMS onset as more commonly done (Ille et al., 2015; Krieg et al., 2017). In the fMRI experiment, the same participants completed three variations of a language paradigm (overt naming, silent naming, and picture observation). The fMRI data were analyzed at three spatial smoothing levels (0, 3, and 6 mm full-width at half-maximum [FWHM]). We then quantified the spatial similarity between the obtained fMRI maps and TMS-induced naming error locations. We expected that task and analysis choices would affect the overlap between fMRI activations and TMS-induced naming error locations (high sensitivity relative to nTMS), while also revealing activations outside error-producing loci (low specificity relative to nTMS), reflecting broader language-related neuronal networks. Concordance between the two modalities was low in general, and depended on the fMRI data smoothing. While the effect of the experimental task remained non-significant, silent naming with 6 mm smoothing yielded the highest group-level similarity to nTMS results. Moreover, within-session fMRI reliability increased with smoothing. These results suggest that analytical choices shape the agreement between TMS- and fMRI-based language mapping, and provide guidance for informing TMS with fMRI and MEG in future studies.

## 2. Methods

### 2.1. Participants

14 healthy, right-handed participants (7 females, ages 22-40, mean age 25.8) participated in the study and gave their written informed consent. The measurement protocol was approved by the Helsinki University Hospital Regional Committee on Medical Research Ethics (HUS/1198/2016). The participants were native Finnish speakers, with no diagnosed neurological disorders, no medication affecting the nervous system, no diagnosed heart conditions, no family history of epilepsy, and no contraindications to MRI or TMS. Normal or corrected-to-normal vision was required, including normal color vision. One participant did not participate in the fMRI part of the study, and another did not participate in the TMS part.

### 2.2. Image set

A single 122-image set was used as stimuli in all measurements (nTMS, fMRI and MEG). The images were retrieved from the bank of standardized stimuli (Brodeur et al., 2010, 2014). The set consisted of color images of everyday objects and included only items with high name agreement and familiarity among native Finnish speakers.

### 2.3. Magnetoencephalography

MEG data was recorded to optimize individual stimulation latencies. The data was recorded with a 306-channel (102 magnetometers, 204 planar gradiometers) whole-head MEG device (Elektra Neuromag TRIUX, Megin Ltd, Helsinki, Finland) in a magnetically shielded room (Euroshield, Eura, Finland), with sampling frequency of 1000 Hz and a bandpass filter of 0.03-330 Hz. The head position of the subject was continuously monitored with 5 head position indicator (HPI) coils. 7 surface electrode pairs were attached to measure heartbeats (electrocardiography), eye movements (electrooculography), and facial muscle activity (electromyography).

The MEG measurement protocol and analysis is described in detail by Tuomaala et al (Tuomaala et al., 2024). During the measurement, the subjects performed a facial gesture task to enable speech artefact removal, and a picture naming paradigm. In the picture naming paradigm, the subjects performed three tasks: overt naming, silent naming, and observation. In the overt naming task, the subjects were shown a picture that they were instructed to name aloud normally and at a brisk pace. In the silent naming task, the subjects were to name a shown picture quietly in their mind, avoiding moving any muscles. In the observation task, the subjects were told to look at the pictures and to keep attention on the task, to identify an occasional overlaying red dot by saying ‘now’. The individual stimulation latencies for the TMS study were obtained from the overt naming task.

### 2.4. Transcranial Magnetic Stimulation

Navigated TMS was delivered using a figure-of-eight coil and a speech mapping system (NBS 5 with NexSpeech, Nexstim, Finland). The navigation system of the stimulator was used to coregister each subject’s T1 MR image to the subject’s head, thus allowing the operator to navigate the TMS system in each subject’s anatomy. At the beginning of the TMS session, the individual TMS pulse intensity was determined based on the resting motor threshold (rMT) of the right hand abductor pollicis brevis (APB) muscle so that 10 out of 20 pulses given to the corresponding M1 area elicited a motor evoked potential of at least 50 μV. Each TMS session started with a baseline naming task of the picture set, repeated two or three times. During the baseline naming, no TMS pulses were given, and any picture that the subject had difficulties to name was removed from the picture set.

The TMS speech mapping protocol followed the guidelines presented in (Lioumis et al., 2023). During the TMS speech mapping, the pictures were displayed for 700 ms with an inter-picture-interval (IPI; timing between onsets of two consecutive images) of 2100-2500 ms (median 2300 ms). The TMS pulse sequence consisted of 5 pulses at a frequency of 5 Hz, except for one subject for which the frequency was increased to 7 Hz to induce more naming errors. By default, the pulse intensity was the same as the rMT (average percent of maximum stimulator output [%MSO] 35.00 ± 3.19 [SEM]). The intensity was decreased if the subject reported pain, and, if tolerable, increased if no naming errors were observed with the initial intensity. Across subjects, the TMS intensity varied from 29 to 40 %MSO (mean 34.35 ± 2.25 [SEM]). The PTI ranged from 0 to 500 ms, including default timings at 0 ms, 200 ms, 300 ms, 400 ms for all except one participant who was not stimulated at 400 ms timing due to tiredness, and various individualized timings based on MEG.

During the cortical speech mapping, the subjects were seated comfortably in a chair. The subjects were instructed to name the shown images at a brisk pace, and in the event of making a naming error, to quickly move on and concentrate on the next image and naming action. Within the measurement, each subject participated in 7–9 runs with varying PTI, with 5–10 minutes break between each run. All runs were monitored with video and audio, using the built-in camera system of the stimulator. To record the naming response times, accelerometer data was successfully recorded for 8 out of 12 subjects using a three-axis accelerometer recorder (ADXL330 iMEMS® Accelerometer, Analog Devices, Norwood, MA) with 3000 Hz sampling frequency attached to the skin either next to or on top of the larynx.

#### 2.4.1. Preprocessing of TMS data

The behavioral TMS data (video and audio recordings of the naming actions) was processed using NexSpeech Analyzer 2.1.0 (Nexstim, Finland). Each naming action was reviewed by a native Finnish speaker by comparing the naming action to the baseline, and then classified either as a naming action with an error, a naming action with no error, or discarded. Naming errors were categorized to five system-defined categories: performance error, no-response error (i.e., anomia), semantic error, other error, or muscle artifact, roughly following the naming error categorization by Corina et al. (Corina et al., 2010). During the error review process, we discarded naming actions during which the TMS coil was moving, was not touching the subject’s head properly, did not stimulate correctly, or if the subject’s naming performance was clearly affected by outside disturbances or actions of the subject not relating to naming. We also discarded naming errors that were caused or affected by a previous error, and repeated semantic errors (the same error repeated at least three times). Naming actions that were not classified as errors nor discarded were classified as non-errors. After classifying all naming actions, the behavioral data was combined with the stimulation data (coil location, orientation, and the estimated electric field magnitude [V/m] at each locus).

The accelerometer data was used to help to identify the naming instances where the speech response time was delayed. The raw accelerometer data was first filtered with a 75 Hz high-pass filter for removing low frequency artifacts. Then, an amplitude envelope of the filtered signal was calculated with a 5-sample moving average to further smooth the resulting signal. From the smoothed envelope, speech onsets were found by locating the first sample where the signal value exceeded 5 μV for at least 150 consecutive samples (5 ms), within epochs spanning from each image onset to the onset of the next image. Delayed responses were classified as performance errors.

Nexstim target coordinates were exported using a peeling depth of 19.3 mm to 23.8 mm (mean 21.4 mm). The locations of maximum electric field within the chosen peeling depth, determined using Nexstim’s spherical head model, were used as TMS target coordinates and transformed to NIfTI voxel space. T1-weighted MRI scans used during TMS were co-registered with T1 scans acquired during fMRI using FSL FLIRT (Jenkinson et al., 2002). NIfTI voxel coordinates were first transformed into millimeter coordinates and then into the same space as the fMRI T1 scans using FLIRT’s *img2stdcoord* utility. Visual inspection was conducted to verify the anatomical correspondence between Nexstim target coordinates and the final co-registered coordinates.

### 2.5. Functional Magnetic Resonance Imaging

#### 2.5.1. fMRI data acquisition

Functional MRI data were acquired using a 3T whole-body MR scanner (Siemens Skyra, 30-channel head-coil) using the CMRR EPI sequence (Moeller et al., 2010) with the following acquisition parameters: TR = 1220 ms, TE = 38 ms, flip angle = 60^°^, multiband factor = 4, 48 slices, 615 volumes, voxel size = 2.6 x 2.6 x 3.12 mm^3^. In order to later correct functional images for geometric distortions using FSL topup (Andersson et al., 2003; Smith et al., 2004), two short EPI measurements were conducted, each with opposing phase-encoding directions. In addition, a T1-weighted image (MPRAGE) was acquired for anatomical reference (TR = 2530 ms, TE = 3.3 ms, flip angle = 7^°^, voxel size = 1 mm^3^ isotropic).

In order to localize language eloquent areas, the participants performed a picture naming task using the same pictures used in the TMS and MEG experiments; see Fig. 1B for a visual summary of the task paradigm. Each picture was projected onto the center of a screen in the bore and displayed for 500 ms. The IPI was set to 3.0 s. The recording consisted of two runs. In the first run, overt naming blocks and picture observation blocks alternated and in the second run overt naming and silent naming blocks alternated. Each task block lasted 30 s, and each condition consisted of 6 blocks of 10 images in each. A rest block of 21 s was presented between each task block. The images were divided into two sets of 60 images that did not differ significantly based on visual complexity (*t*_115_ = 0.016, *P* = 0.99), word length (*t*_120_ = −0.31, *P* = 0.76) or word frequency (*t*_120_ = 0.35, *P* = 0.72). Different image sets were presented during the two conditions within a run. During the second run, the same images were reused with scrambled set assignment: overt naming received half from its own first-run set and half from the observation set, and silent naming received the complementary halves.

**Figure 1:**
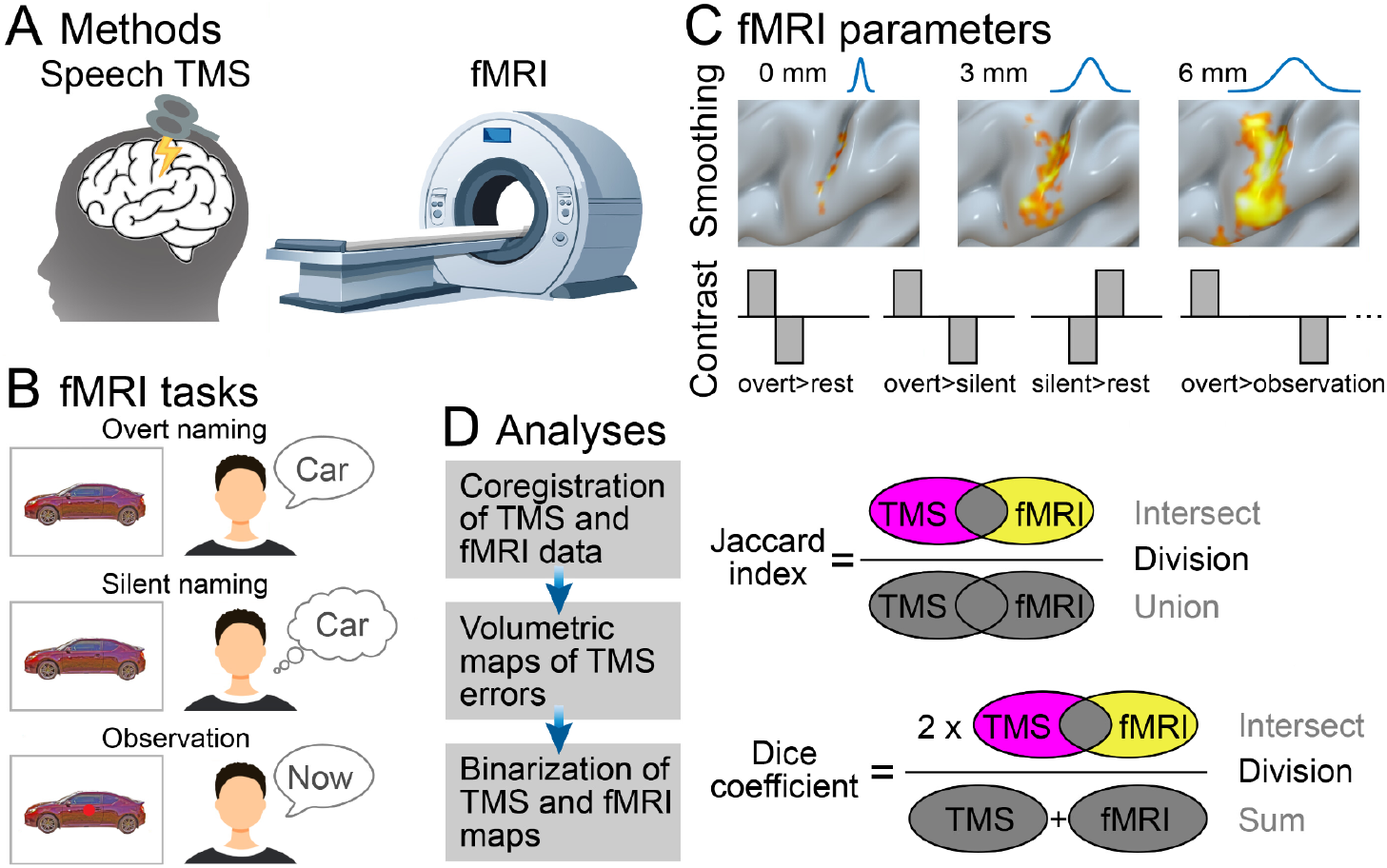
Schematic description of the study. A) Methods used in the current study. B) Tasks used in the fMRI experiment. C) fMRI parameters included three smoothing levels and several task contrasts (only four of the used contrasts shown as examples). D) Analyses included Euclidean distance calculations between TMS locations and fMRI cluster peaks. Jaccard index was used to quantify the similarity between TMS and fMRI data.

In the overt naming task, the subjects were shown a picture that they were instructed to name aloud, while in the silent naming task, the subjects were asked to name a shown picture quietly in their mind. In the observation task, the subjects were told to look at the pictures and to identify an occasional overlaying red dot by saying ‘now’ (25% of the images); the task instructions were similar in MEG and fMRI. During the rest block (12 blocks in total), subjects were instructed to look at a fixation cross. The total task runtime was 12 min and 50 s.

#### 2.5.2 fMRI preprocessing

fMRI data were processed using a pipeline that integrated tools from FSL (Jenkinson et al., 2012) and SPM12 (Statistical Parametric Mapping, https://www.fil.ion.ucl.ac.uk/spm/). First, the DICOM files were converted to NIfTI format using *dcm2niix* (Li et al., 2016). To correct for geometric distortions, we utilized FSL’s *topup* tool, employing the two EPI images with opposing phase-encoding directions (Andersson et al., 2003). Motion artifacts were addressed using SPM12’s realignment by estimating the motion parameters and reslicing the images to the mean functional image. Framewise displacement was calculated to identify motion artifacts, with a 3-mm threshold set as a criterion for excluding data (Power et al., 2012). No datasets exceeded this threshold, indicating that all data passed the quality check (Fig. S1-2). The functional images were then coregistered to each participant’s structural T1-weighted image. To conduct group-level analyses, the realigned and coregistered images were additionally normalized to MNI space. Spatial smoothing of the functional images was performed using a Gaussian kernel with three different smoothing levels: FWHM of 0 mm, 3 mm and 6 mm.

#### 2.5.3. fMRI first-level analysis

Single-subject (first-level) analysis was performed using a general linear model. Linear regression was performed for each voxel based on generalized least squares with a global approximate AR(1) autoregression model, drift-fit with Discrete Cosine Transform basis (128 s cutoff). Task blocks of 30 s each and baseline blocks of 21 s were convolved with SPM’s canonical HRF and used as regressors of interest. Realignment parameters obtained from the previous preprocessing steps were included in the model as nuisance regressors. A binary brain mask was used to exclude voxels outside the brain. Single-subject *t*-maps were calculated in subject native space for each contrast. The threshold of the *t*-statistic was set to *P*<0.001, with cluster-wise FWE correction (initial cluster defining threshold <0.001; see Table S1-2 for k extent thresholds).

To conduct group-level (second-level) analysis, the re-sliced and coregistered data was normalized to MNI space using 4th Degree B-Spline interpolation and 2 mm isotropic voxel size. Then, the normalized data was smoothed using FWHM of 0 mm, 3 mm and 6 mm. The fist-level analyses were also conducted for MNI-normalized single subject data with otherwise same parameters as the native space data except that no masking was applied.

#### 2.5.4. fMRI second-level analysis

For the group-level fMRI analysis, a one-sample t-test design was specified, incorporating all subjects for each contrast and smoothing level. Parameter estimates were calculated, and *t*-maps were generated using an initial cluster-defining threshold of *P*<0.001 (uncorrected) and cluster-level FWE correction.

#### 2.5.5. fMRI reliability

The overt object naming task was repeated twice per fMRI scanning session, separated in two scanning runs (∼22 min between the two runs). Thus, we were able to quantify the within-session reliability of the naming>rest condition. Unthresholded, spatially normalized 1st level T-maps were used to quantify concordance correlation coefficient (CCC; (Lin, 1989)) for each voxel. The resulting CCC maps were thresholded for visualization purposes. In addition to voxelwise CCC maps, we also calculated subjectwise CCC values (reliability across voxels within all subjects).

### 2.6. Quantifying the similarity between TMS and fMRI data

To compare the similarity across TMS and fMRI mapping results within each subject, we first created volumetric density maps for both the TMS locations producing naming errors and for those not producing naming errors. We applied a 3-D Gaussian smoothing kernel (1 mm sigma) to each TMS coordinate and summed these Gaussian fields across all stimulation sites to form density maps. These maps were normalized by dividing all voxel values by its maximum value and subsequently binarized. The error-specific density maps were then created by subtracting the non-error density map from the error density map, ensuring that all negative values were set to zero and thus emphasizing the regions corresponding to error specificity. In addition to Jaccard indices, we also calculated naming error proximity indices for each comparison (Fig. S4).

Binary masks were created by summing up 10 mm spherical areas around all TMS spots of a given subject. Within these masks, Jaccard indices were calculated as the ratio of the number of voxels common to both the TMS error-specific density and the binarized fMRI T-maps to the total number of voxels present in either map (intersection over union). In addition to Jaccard indices, we also calculated the Dice coefficients for each comparison, defined as twice the number of voxels in the intersection divided by the sum of the voxel counts in the two maps (Fig. S3).

In addition to the native space calculations, we also performed an additional, group-level analysis to compare similarity between TMS and fMRI mapping. For that purpose, we normalized the error-specific density maps into MNI space and created a mean error density map using SPM’s *imcalc* (trilinear interpolation). Jaccard indices were then calculated using binarized, 2nd level fMRI maps.

### 2.7. Statistical analyses

To test the effect of fMRI data smoothing and contrast on Jaccard indices and Dice coefficients, we performed two-way repeated-measures analyses of variance (rmANOVAs) with Geisser-Greenhouse correction. The effect of data smoothing on subjectwise CCC was tested using a one-way rmANOVA, followed by post hoc Tukey’s tests.

## 3. Results

### 3.1. TMS results

All subjects responded to the left-hemispheric TMS stimulation by making speech errors during the overt naming task. The number of successful stimuli given during the experiment was on average 1153 per subject (range 893 to 1381). On average, 37 speech errors (range 9 to 75) were made per subject during the whole experiment, corresponding to an average error rate of 3.2% (range 0.75% to 7.4%); the error rate was roughly in line with earlier studies in healthy adults (Picht et al., 2013). At the group level, a total of 443 speech errors were recorded. 66.1% of these were performance errors (delayed, stuttered and slurred speech), 11.6% anomias, 7.0% semantic errors, and 15.9% other errors (mostly phonological paraphasias). Spatial distributions of nTMS-induced naming errors also corresponded with earlier studies, showing the largest amount of error in frontal and parietal areas (Fig. 2 and Supplementary Material; (Ille et al., 2015; Picht et al., 2013)).

**Figure 2:**
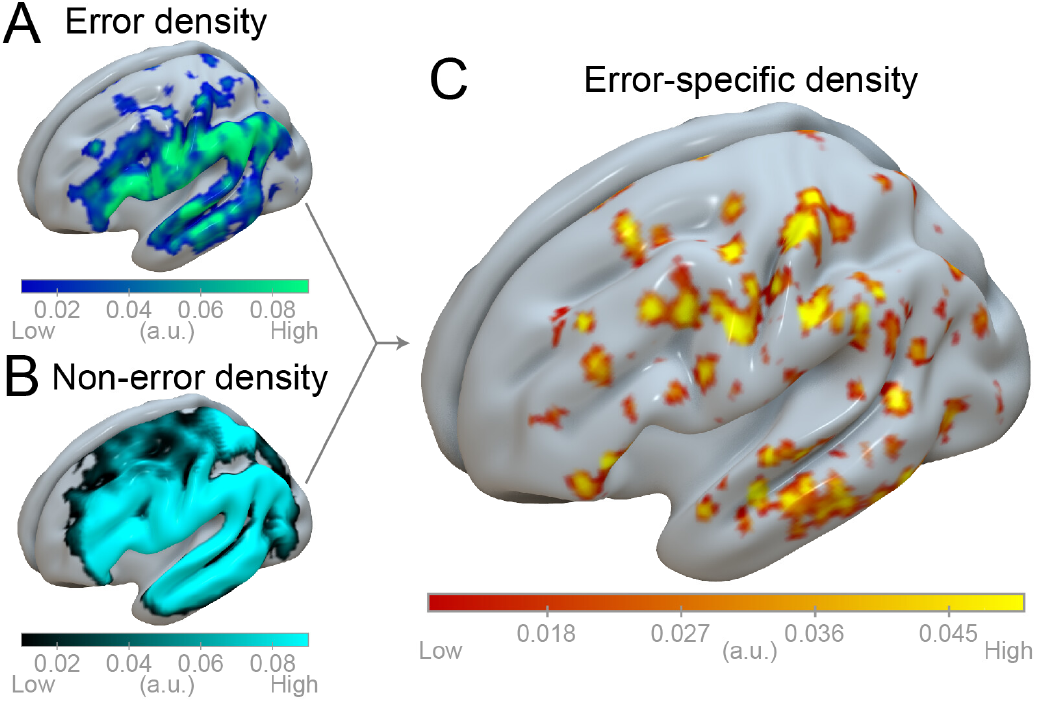
TMS group-level results (N=12). A) TMS speech error density. B) TMS non-error density. C) TMS naming-error-specific density. The density maps are thresholded at >0.01 (a.u.) for illustratory purposes.

### 3.2. fMRI results

To assess how fMRI parameters affect language mapping sensitivity within subjects, we created 1st level, native space T-maps for each contrast and smoothing level. Across contrasts and subjects, the number of voxels surviving the thresholding increased from 3425 ± 336 (SEM) with 0 mm smoothing, to 7052 ± 669 with 3 mm smoothing and 10208 ± 996 with 6 mm smoothing. Mean T-value of the significantly activated voxels was 4.93 ± 0.056 with 0 mm smoothing, 5.06 ± 0.076 with 3 mm smoothing, and 5.14 ± 0.091 with 6 mm smoothing, respectively.

In addition, we ran a 2nd level fMRI analysis (Fig. 3). Across contrasts, the number of significant clusters decreased from 7.72 (range 1 to 18) for 0 mm smoothing, to 5.37 (range 1 to 13) for 3 mm smoothing, and to 4.29 (range 1 to 10) for 6 mm smoothing. The average cluster sizes increased from 40.47 (range 12 to 153) to 324.90 (range 30 to 4386) and to 1035.09 (range 106 to 10074), respectively. Mean T-value of the significantly activated voxels was 6.77 ± 0.14 (0 mm smoothing), 7.82 ± 0.33 (3 mm), and 7.87 ± 0.42 (6 mm). To map the activated regions to anatomical structures, we utilized the AAL3 atlas (Rolls et al., 2020). Table S3 highlights the predominant anatomical locations of the largest significant clusters, along with peak voxel coordinates within those clusters.

**Figure 3:**
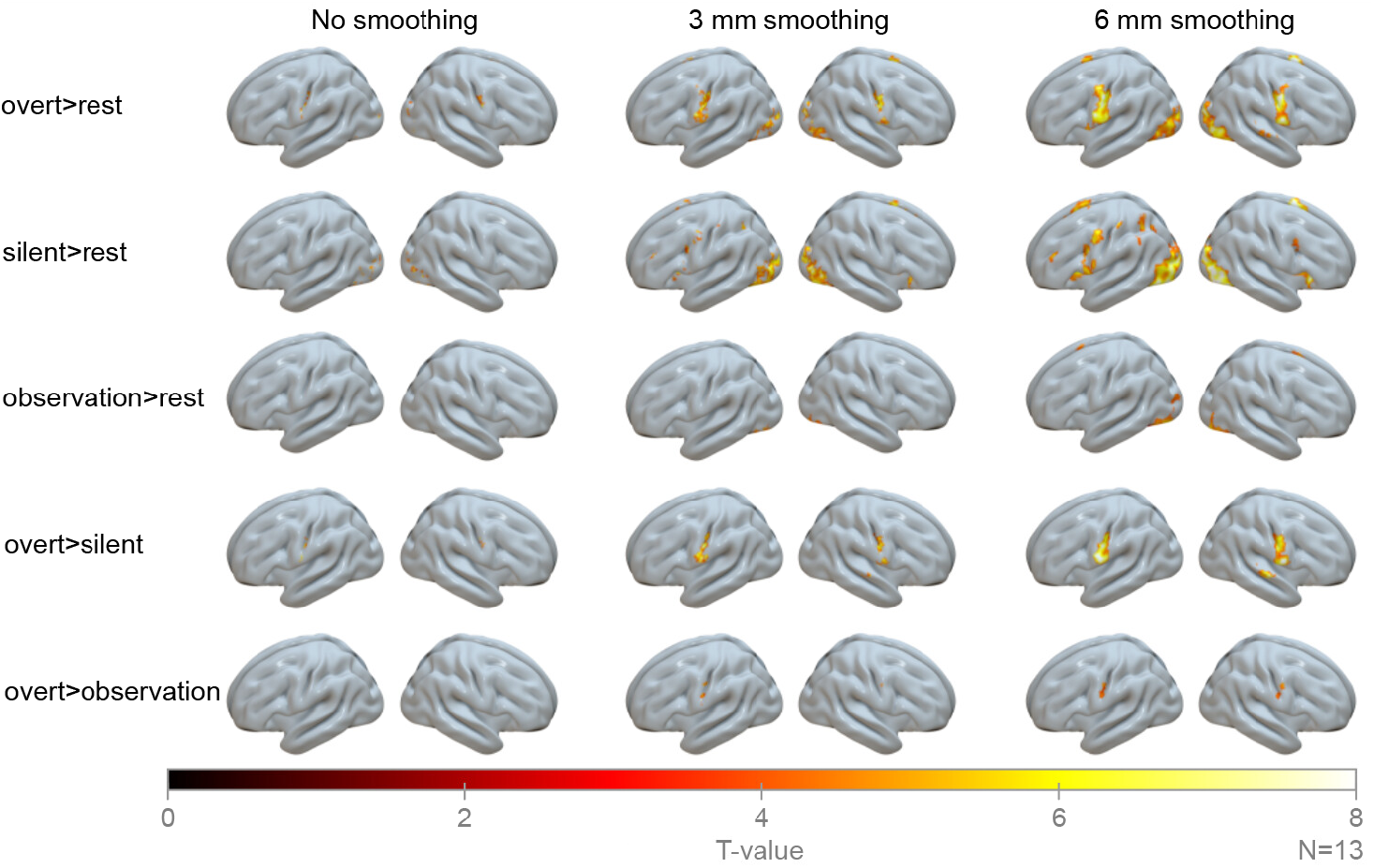
2nd level fMRI results. 2nd level results for each contrast and smoothing level (height threshold: *P*<0.001 (uncor.); FWE cluster extent threshold; N=13).

### 3.3. Comparison between TMS and fMRI mapping

We quantified the spatial correspondence between TMS and fMRI by computing Jaccard indices within each subject between binarized fMRI activation maps and TMS-derived error-specific density maps, across contrasts and smoothing levels (Fig. 4). Overall, the within-subject similarity of the maps was low: the mean error-specific Jaccard index across all smoothing levels, contrasts, and subjects was 0.036 ± 0.0086. Simple overlap counts between fMRI and TMS data were higher for non-error (1496 voxels) than error locations (732 voxels), likely reflecting the larger number of non-error sites. The overlap with error-specific locations was 115 voxels. Smoothing influenced the map similarity: for error-specific maps, all but one subject showed their maximum Jaccard index at 6 mm FWHM (one subject at 0 mm); for error density maps, all subjects had their maxima at 6 mm. The experimental contrast that maximized the correspondence between TMS and fMRI (‘optimal’ contrast) varied across individuals (Table 1). At the group level (Fig. 4E–F), the condition yielding the highest error-specific Jaccard index was silent naming>rest with 6 mm smoothing (mean ± SEM 0.053 ± 0.014). A two-way rmANOVA of the Jaccard indices showed a significant main effect of smoothing (*F*_2,22_ = 15.15, *P* = 0.0020), but a non-significant main effect of contrast (*F*_4,44_ = 2.46, *P* = 0.11) and a non-significant interaction of smoothing and contrast (*F*_8,88_ = 0.45, *P* = 0.72). Two-way rmANOVA of the Dice coefficients was in line with Jaccard index analyses: main effect of smoothing was statistically significant (*F*_2,22_ = 13.36, *P* = 0.0032) whereas the main effect of contrast (*F*_4,44_ = 2.58, *P* = 0.10) and the interaction of smoothing and contrast (*F*_8,88_ = 0.44, *P* = 0.72) were not.

**Table 1:** Optimal contrasts within individuals. The experimental contrast that maximized TMS vs. fMRI concordance (‘optimal’ contrast, quantified with Jaccard index) varied across individuals. Each cell in the table shows the amount of participants having that contrast as the optimal, both for error-specific density and error density maps.

|  | Experimental contrast |  |  |  |  |
| --- | --- | --- | --- | --- | --- |
|  | overt>rest | silent>rest | observation>rest | overt>silent | overt>observation |
| Error-specific density | 2/12 | 4/12 | 3/12 | 2/12 | 1/12 |
| Error density | 2/12 | 3/12 | 3/12 | 1/12 | 3/12 |

**Figure 4:**
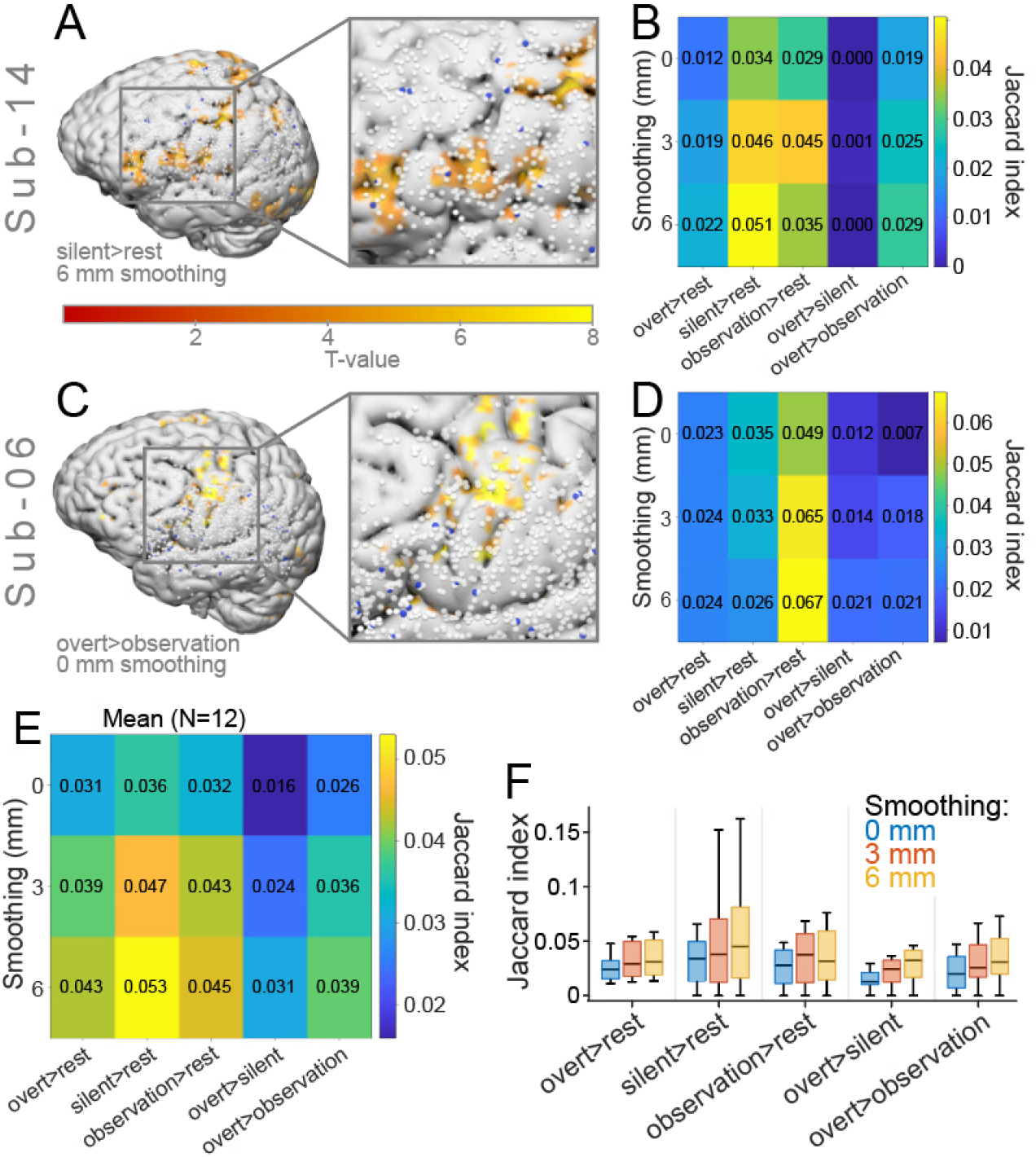
Comparison of TMS and fMRI mapping data in native space. A) One participant’s (silent naming>rest, 6 mm smoothing) TMS and fMRI mapping data with a magnified view, and a heatmap showing Jaccard indices between TMS and fMRI mapping data for each fMRI contrast and smoothing level (B). C) One participant’s (overt naming>observation, 0 mm smoothing) TMS and fMRI mapping data with a magnified view and a heatmap showing Jaccard indices between TMS and fMRI mapping data in each fMRI contrast and smoothing level (D). E-F) Group-average (N=12) heatmap (E) and boxplot (F) showing averaged Jaccard indices for each fMRI contrast and smoothing level. Boxes in F represent median and interquartile range, and the whiskers denote the minima and maxima.

In addition to the within-subject comparisons, we quantified Jaccard indices between the group-level TMS error-specific density maps and the second-level fMRI maps (Fig. 5). The Jaccard index across all contrasts and smoothing levels was low, 0.0071 ± 0.00067 (mean ± SEM). By smoothing level, the mean Jaccard indices were 0.00074 (0 mm), 0.0057 (3 mm), and 0.0148 (6 mm). Among contrasts, the mean Jaccard indices were 0.013 (overt naming>rest), 0.0160 (silent naming>rest), 0.0022 (observation>rest), 0.0034 (overt naming>silent naming), and 0.0012 (overt naming>observation). The largest Jaccard index was observed for silent naming>rest combined with 6 mm smoothing. Dice coefficients showed the same pattern across contrasts and smoothing levels as the Jaccard indices (Fig. S3).

**Figure 5:**
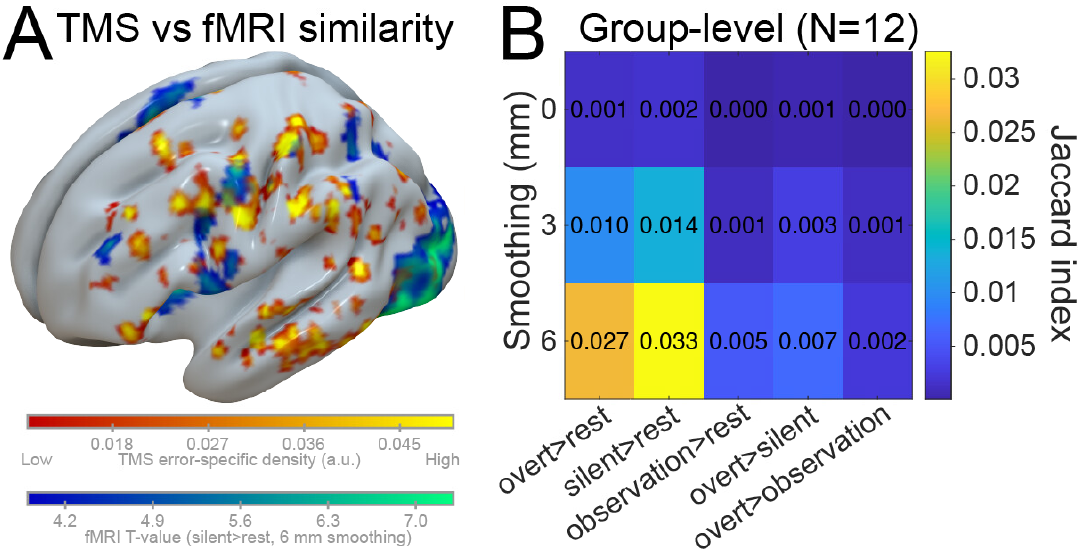
TMS vs. fMRI: Group-level comparisons. A) TMS naming-error-specific density overlaid with silent naming>rest 2nd level map (6 mm smoothing). B) The heatmap shows Jaccard similarity indices between group-level TMS naming-error-specific density and 2nd level fMRI maps across smoothing levels and contrasts (N=12).

### 3.4. fMRI reliability

To evaluate the test-retest reliability of fMRI results, we ran the overt naming task twice per session, and quantified CCC for each voxel (Fig. 6). Voxelwise CCC increased with the level of smoothing: median CCC_0mm_ = 0.42 (interquartile range [IQR] 0.35), median CCC_3mm_ = 0.47 (IQR 0.33) and median CCC_6mm_ = 0.50 (IQR 0.29; Fig. 6). In line with that, rmANOVA showed that subjectwise CCC differed significantly across smoothing levels (*F*_2,24_ = 17.29, *P* = 0.0012). Post hoc testing showed significant differences between 0 mm versus 3 mm (*P* = 0.0015), 0 mm versus 6 mm (*P* = 0.0036), and 3 versus 6 mm smoothing (*P* = 0.012). Taken together, a larger amount of smoothing increased the test-retest reliability.

**Figure 6:**
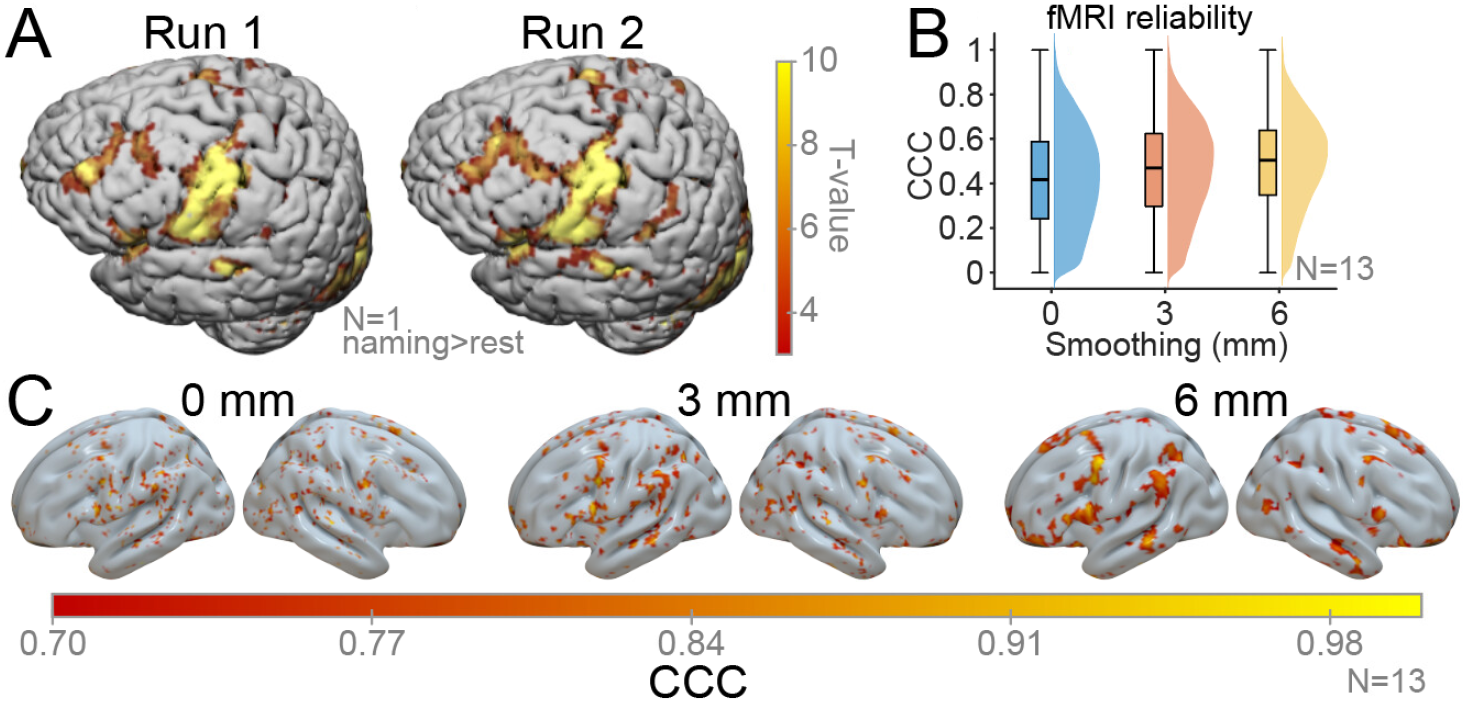
The effect of smoothing on fMRI reliability. A) Example of one subject’s overt naming>rest contrast across two runs (6 mm smoothing, FWE cluster extent threshold). B) Voxelwise concordance correlation coefficient (CCC) across smoothing levels for overt naming>rest contrast. The bars and violins represent all voxels (N=13). C) Voxelwise CCC across smoothing levels for overt naming>rest contrast (N=13). CCC was calculated using non-thresholded T-maps, but for visualization purposes the figures here are thresholded at CCC>0.7 and clusters <20 voxels have been removed.

## 4. Discussion

### 4.1. Summary of the findings

We examined how fMRI task choice and extent of spatial smoothing affect concordance with nTMS SCM. Across 12 healthy volunteers, we combined individualized nTMS during overt naming with three language tasks conducted with fMRI (overt naming, silent naming, and image observation) analyzed at 0, 3, and 6 mm FWHM smoothing. Concordance between modalities was low overall, and depended on fMRI data smoothing. The effect of experimental task on fMRI versus nTMS concordance was statistically non-significant, but silent naming with 6 mm smoothing yielded the highest overlap with nTMS error-specific maps at single-subject and group levels. Within-session fMRI reliability increased with greater smoothing.

Earlier studies have emphasized task dependence and methodological choices in presurgical language mapping and noted divergence across modalities that are likely to probe different aspects of language processing (e.g., fMRI activation versus nTMS or DCS perturbation; (Hauck et al., 2019; Ille et al., 2015; Sollmann et al., 2016; Tarapore & Nagarajan, 2017)). Compared with earlier studies, we here sought to minimize possible sources of bias and confounds by using distortion-corrected fMRI (Andersson et al., 2003), data-driven concordance mapping without a priori parcels, and multiple objective overlap metrics (Jaccard, Dice, and proximity indices). Furthermore, we used both fixed and MEG-based, personalized TMS latencies, and systematic manipulation of spatial smoothing of the fMRI data. Our results support earlier results on the complementary nature of nTMS and fMRI and suggest that if integrating fMRI with MEG-informed nTMS for speech mapping, silent naming with moderate smoothing (≈6 mm) may modestly improve alignment, but low absolute overlap should be expected.

### 4.2. MEG-informed nTMS: error profiles and timing

We conducted individualized, MEG-informed nTMS SCM of the left hemisphere using multiple picture-to-TMS intervals. nTMS induced speech errors in all participants, with a mean error rate of 3.2% across an average of 1155 valid stimuli per subject. The error types and their distributions are consistent with studies demonstrating that nTMS can elicit transient speech disruptions and that error profiles and spatial distributions vary across individuals and tasks (Krieg et al., 2017; Lioumis et al., 2012). Relative to DCS, nTMS has been reported to be sensitive but less specific (Picht et al., 2013), emphasizing the need for optimizing timing and sampling density to balance false positive and negative findings (Jeltema et al., 2021). Future work should explore more the sampling density and incorporate individualized MEG-informed timings to improve the efficiency and interpretability of nTMS SCM.

### 4.3. fMRI SCM: reliability and effects of smoothing

Reliability (voxel- and subjectwise CCC) of fMRI results across two runs of the overt naming task slightly increased with greater smoothing (median voxelwise CCC rising from 0.42 at 0 mm to 0.50 at 6 mm), illustrating the trade-off between spatial precision and robustness. This pattern aligns with reports that spatial smoothing can increase intraindividual reproducibility (Ning et al., 2019; Vasileiadi, Woletz, et al., 2023). At the same time, clinical fMRI SCM work has shown that fMRI is prone to false positive findings (Brennan et al., 2016; Leone et al., 2025), underscoring the limits on single-subject precision. In future work, it would be valuable to explore smoothing levels beyond 6 mm, assess test-retest reliability across tasks, and explicitly test how other preprocessing choices (e.g., distortion correction) affect reliability and concordance with nTMS- and DCS-based mapping.

### 4.4. Concordance between nTMS and fMRI SCM

To evaluate the concordance between nTMS- and fMRI-based SCM, we constructed nTMS-induced-error-specific volumetric maps, and compared them to binarized fMRI maps using Jaccard and Dice indices. Within-subject spatial overlap was low overall (mean error-specific Jaccard index 0.036). Smoothing influenced the concordance: most participants showed maximal Jaccard at 6 mm FWHM. The optimal contrast varied across individuals, but at the group level, silent naming>rest contrast with 6 mm smoothing yielded the highest overlap between the modalities. These results are consistent with prior studies reporting limited agreement between fMRI activation and nTMS language-positive loci, task dependencies, and regional variability (Hauck et al., 2019; Schiller et al., 2020b; Sollmann et al., 2016). Reported comparisons to DCS also indicate that nTMS often shows closer correspondence to DCS than fMRI, especially for motor mapping but also in some SCM studies (Babajani-Feremi et al., 2016; Coburger et al., 2013; Forster et al., 2011; Muir et al., 2022; Picht et al., 2013). While the earlier studies suggested that the spatial distributions across the two modalities would differ, the Jaccard indices seen in the current study were lower than we initially expected. We interpret that nTMS pinpoints causal nodes where transient disruption produces naming errors, while fMRI reflects broader task-engaged networks, leading to low overlap at the voxel level. Potential contributors to residual disagreement include task differences in silent naming and observation tasks, regional susceptibility and possible distortion especially in the temporal lobe, and operator-dependance in TMS sampling density and error classification. Another possible explanation for higher fMRI vs. TMS concordance in silent naming compared to overt naming task could be that the primary motor activation dominates the fMRI response in the overt condition, masking possible language-related activations. The current data-driven approach (error-specific density rather than atlas-based parcels) offers finer granularity than earlier parcel-wise comparisons. Future work should test additional language tasks known to elicit different error profiles in nTMS (Hauck et al., 2015), and prospectively evaluate the use of fMRI to guide nTMS sampling density. In conclusion, our data-driven overlap metrics, individualized nTMS timing, and systematic manipulation of task and smoothing provide quantitative benchmarks and practical parameters (e.g., silent naming with 6 mm FWHM) for designing integrated fMRI-nTMS SCM paradigms, and a reproducible framework to test further refinements.

### 4.5. Limitations and future directions

Several strengths enhance the rigor and reproducibility of the current study: distortion-corrected fMRI, within-session reliability quantification, MEG-informed individualized picture-to-TMS intervals, objective overlap metrics, a systematic manipulation of spatial smoothing across tasks, and a dense nTMS dataset per participant (mean 1155 stimuli) analyzed with a data-driven error-specific approach. Nevertheless, there are limitations to consider. The sample size was modest, and residual susceptibility/distortion may persist in temporal regions despite correction. While effective stimulation locations were estimated with realistic e-field modeling, the current paradigm has been shown to be clinically robust. Future work should increase sample size, evaluate test-retest reliability of nTMS SCM, broaden the task set (e.g., pseudoword reading, verb generation, action naming), incorporate individualized fMRI-informed targeting prospectively, and add realistic e-field modeling. Integrating fMRI and MEG to guide both where and when to stimulate (Ala-Salomäki et al., 2021; Grummich et al., 2006; Könönen et al., 2015; Tarapore & Nagarajan, 2017) remains a feasible path to personalize nTMS timing and sampling uncertainties in SCM.

## 5. Conclusions

In the current study, we evaluated spatial concordance between nTMS and task-based fMRI for mapping language-critical areas in the left hemisphere. Concordance between modalities was overall very low and systematically influenced by data smoothing choices. Across individuals and at the group level, the silent naming>rest contrast with 6 mm FWHM smoothing yielded the highest overlap with nTMS maps, while greater smoothing modestly improved within-session fMRI reliability. These findings are consistent with the complementary nature of perturbation-based (nTMS) and hemodynamic (fMRI) measures and indicate that analytical parameters shape their agreement. Practically, in the case of integrating fMRI with individualized nTMS SCM, silent naming with moderate smoothing may enhance alignment, though absolute overlap remains limited and variable across individuals. Our framework provides quantitative benchmarks for multimodal language mapping and a basis for prospective optimization of task design, and analytical strategies.

## Data Availability

Data produced in the present study are available upon reasonable request to the authors

## CRediT authorship contribution statement

**JG**: Conceptualization, Methodology, Software, Validation, Formal analysis, Investigation, Data Curation, Writing - Original Draft, Writing - Review & Editing, Visualization, Project administration, Funding acquisition. **SA:** Conceptualization, Methodology, Software, Validation, Formal analysis, Investigation, Data Curation, Writing - Original Draft, Writing - Review & Editing, Project administration. **MV**: Conceptualization, Methodology, Formal analysis, Writing - Review & Editing. **MT**: Methodology, Writing - Review & Editing, Funding acquisition. **SV**: Conceptualization, Methodology, Writing - Review & Editing, Funding acquisition. **HR**: Conceptualization, Methodology, Resources, Writing - Review & Editing, Supervision, Project administration, Funding acquisition. **ML**: Conceptualization, Methodology, Software, Validation, Data Curation, Writing - Review & Editing, Supervision, Project administration. **PL**: Conceptualization, Methodology, Investigation, Resources, Data Curation, Writing - Review & Editing, Supervision, Project administration, Funding acquisition.

## Declaration of Interest

SV has been working as an end-user device tester in Nexstim Plc. PL has been a consultant for Nexstim Plc. for speech mapping purposes. All other authors have nothing to disclose.

## Acknowledgements

The study was funded by HUS VTR (TYH2022224); Research Council Finland (357660 to PL and SV, UAK780VAAL, 355409 to HR, and the Flagship of Advanced Mathematics for Sensing, Imaging, and Modeling grant 359181 to HR); the Sohlberg Foundation (to HR); Maire Taposen säätiö (to PL); and the Emil Aaltonen Foundation (personal grant to JG). We thank the study participants and, in particular, Sara Korkealaakso, née Tuomaala, who measured and preprocessed the picture naming MEG data that supported this work.

## Notes

### Funding Statement

HUS VTR (TYH2022224); Research Council Finland (357660, UAK780VAAL, 355409 to HR and Flagship of Advanced Mathematics for Sensing, Imaging, and Modeling grant 359181 to HR); Sohlberg Foundation (to HR); Maire Taponen foundation (to PL); Emil Aaltonen Foundation (personal grant to JG)

### Author Declarations

The study protocol was approved by the Helsinki University Hospital Regional Committee on Medical Research Ethics (HUS/1198/2016).

