## Supplementary Material for "fMRI analysis parameters affect the concordance with TMS in noninvasive speech mapping"

Gogulski J*δ[1,2], Autti S*[1,3], Vasileiadi M[4,5], Tik M[5], Vaalto S[2], Renvall H[1,3], Liljeström M*[1,3], Lioumis P*[1,3, 6]

**Affiliations:**
1 Department of Neuroscience and Biomedical Engineering, Aalto University School of Science, Espoo, Finland
2 Department of Clinical Neurophysiology, HUS Diagnostic Center, Helsinki University Hospital and University of Helsinki, Helsinki, Finland
3 BioMag Laboratory, HUS Diagnostic Center, Helsinki University Hospital, University of Helsinki and Aalto University School of Science, Helsinki, Finland
4 Sunnybrook Health Sciences Center, University of Toronto, Toronto, Ontario, Canada
5 Medical University of Vienna, Vienna, Austria
6 Cognitive Brain Research Unit, Department of Psychology and Logopedics, Faculty of Medicine, University of Helsinki, Helsinki, Finland.

***Equal contribution**

**δCorrespondence**

*Spatial distributions of different nTMS-induced naming error types*

Overall, 43.8% of the nTMS-induced naming errors were observed after stimulation of the parietal lobe, 32.7% of the frontal lobe, and 22.3% of the temporal lobe. 1.1% of the errors were observed after stimulation of the occipital lobe which was not a primary target stimulation area in the current study. No-response errors and other error types were present in roughly equal proportions in the frontal, temporal, and parietal lobes (37.3%, 17.6%, 45.1% of no-response errors, and 35.7%, 17.1%, 47.1% of other errors). Compared to no-response and other errors, performance errors and semantic errors were relatively more frequent after stimulation of the temporal lobe (23.4% of performance errors and 32.3% of semantic errors) and relatively less present in the frontal lobe (31.6% of performance errors and 29.0% of semantic errors). In addition, we recorded 21 instances of speech error-like behaviour (10 after frontal lobe and 11 after temporal lobe stimulation), which we classified as errors made due to muscle stimulation: these errors were not included in the analyses.

| | No smoothing | | | | | | | | | | | | 3 mm smoothing | | | | | | | | | | | | 6 mm smoothing | | | | | | | | | | | |
|---|---|---|---|---|---|---|---|---|---|---|---|---|---|---|---|---|---|---|---|---|---|---|---|---|---|---|---|---|---|---|---|---|---|---|---|---|
| | Contrast number | | | | | | | | | | | | Contrast number | | | | | | | | | | | | Contrast number | | | | | | | | | | | |
| **Sub** | **1** | **2** | **3** | **4** | **5** | **6** | **7** | **8** | **9** | **10** | **11** | **12** | **1** | **2** | **3** | **4** | **5** | **6** | **7** | **8** | **9** | **10** | **11** | **12** | **1** | **2** | **3** | **4** | **5** | **6** | **7** | **8** | **9** | **10** | **11** | **12** |
| 2 | 8 | 8 | 8 | 8 | 8 | 8 | 8 | 8 | 8 | 8 | 8 | 8 | 15 | 15 | 15 | 20 | 15 | 16 | 15 | 20 | 15 | - | 16 | 15 | 50 | 54 | 53 | 41171 | 69 | 56 | 63 | 71 | 108 | 41548 | 51 | 60 |
| 3 | 7 | 7 | 7 | 7 | 7 | 7 | 7 | 7 | 7 | 7 | 7 | 7 | 16 | 17 | 16 | 24 | 30 | 17 | 16 | 16 | - | - | 17 | 16 | 89 | 69 | 54 | 43869 | 53 | 55 | 76 | 125 | 49473 | 42176 | 73 | 53 |
| 4 | 7 | 7 | 7 | 7 | 7 | 7 | 7 | 7 | 7 | 7 | 7 | 7 | 15 | 16 | 16 | 15 | 15 | 15 | 15 | 15 | 15 | 20 | 17 | 15 | 82 | 54 | 48 | 63 | 63 | 51 | 50 | 69 | 61 | 100 | 98 | 50 |
| 5 | 7 | 7 | 7 | 7 | 7 | 7 | 7 | 7 | 7 | 7 | 7 | 7 | 15 | 17 | 19 | 18 | 20 | 15 | 18 | 17 | 15 | 15 | 15 | 16 | 283 | 44 | 53 | 52 | 49 | 44 | 38762 | 61 | 45 | 57 | 44 | 96 |
| 6 | 6 | 6 | 6 | 6 | 6 | 6 | 6 | 6 | 6 | 6 | 6 | 6 | 14 | 17 | 14 | 14 | 14 | 14 | 14 | 14 | 14 | 14 | 14 | 15 | 42 | 110 | 41 | 45 | 46 | 58 | 43 | 42 | 41 | 44 | 45 | 55 |
| 7 | 6 | 6 | 6 | 6 | 6 | 6 | 6 | 6 | 6 | 6 | 6 | 6 | 15 | 14 | 14 | 14 | 14 | 14 | 15 | 14 | 14 | 14 | 14 | 14 | 51 | 57 | 45 | 49 | 84 | 45 | 56 | 67 | 65 | 46 | 55 | 53 |
| 8 | 7 | 7 | 7 | 7 | 7 | 7 | 7 | 7 | 7 | 7 | 7 | 7 | 15 | 20 | 15 | 16 | 17 | 16 | 18 | 24 | 15 | 15 | 15 | 15 | 54 | 48 | 129 | 70 | 47 | 99 | 53 | 59 | 49 | 46 | 53 | 48 |
| 10 | 6 | 6 | 6 | 6 | 6 | 6 | 6 | 6 | 6 | 6 | 6 | 6 | 14 | 14 | 14 | 14 | 15 | 14 | 14 | 14 | 14 | 15 | 14 | 18 | 47 | 45 | 43 | 43 | 57 | 55 | 44 | 44 | 42 | 42 | 42 | 57 |
| 11 | 6 | 6 | 6 | 6 | 6 | 6 | 6 | 6 | 6 | 6 | 6 | 6 | 14 | 13 | 13 | 13 | 14 | 13 | 13 | 13 | 13 | 13 | 13 | 14 | 51 | 40 | 39 | 48 | 40 | 48 | 50 | 359 | 63 | 45 | 66 | 101 |
| 12 | - | 6 | 6 | 6 | 6 | 6 | 6 | 6 | 6 | 6 | 6 | 6 | - | 14 | 14 | 14 | 14 | 16 | 15 | 25 | 15 | 16 | 15 | 15 | - | 42 | 88 | 45 | 42 | 46 | 56 | 62 | 56 | 72 | 46 | 50 |
| 13 | 6 | 6 | 6 | 6 | 6 | 6 | 6 | 6 | 6 | 6 | 6 | 6 | 14 | 14 | 14 | 15 | 24 | 14 | 14 | 14 | 14 | 14 | 15 | 14 | 57 | 43 | 74 | 46 | 47 | 59 | 49 | 45 | 70 | 92 | 57 | 92 |
| 14 | 6 | 6 | 6 | 6 | 6 | 6 | 6 | 6 | 6 | 6 | 6 | 6 | 14 | 14 | 14 | 14 | 14 | 14 | 15 | 15 | 14 | 15 | 14 | 14 | 60 | 42 | 41 | 46 | 42 | 59 | 44 | 42 | 52 | 49 | 149 | 58 |
| 15 | 6 | 6 | 6 | 6 | 6 | 6 | 7 | 6 | 6 | 6 | 6 | 6 | 13 | 13 | 15 | 13 | 15 | 13 | 15 | 15 | 15 | 13 | 13 | 13 | 50 | 41 | 40 | 53 | 42 | 42 | 40 | 45 | 51 | 41 | 138 | 41 |

**Table S1 - Cluster extent thresholds for 1st level, native space data of the first fMRI run.** Conditions in which there were no significant clusters are marked with ‘-’.

| | No smoothing | | | | | | | | | | | | 3 mm smoothing | | | | | | | | | | | | 6 mm smoothing | | | | | | | | | | | |
|---|---|---|---|---|---|---|---|---|---|---|---|---|---|---|---|---|---|---|---|---|---|---|---|---|---|---|---|---|---|---|---|---|---|---|---|---|
| | Contrast number | | | | | | | | | | | | Contrast number | | | | | | | | | | | | Contrast number | | | | | | | | | | | |
| **Sub** | **1** | **2** | **3** | **4** | **5** | **6** | **7** | **8** | **9** | **10** | **11** | **12** | **1** | **2** | **3** | **4** | **5** | **6** | **7** | **8** | **9** | **10** | **11** | **12** | **1** | **2** | **3** | **4** | **5** | **6** | **7** | **8** | **9** | **10** | **11** | **12** |
| 2 | 6 | 6 | 6 | 6 | 6 | 6 | 6 | 6 | 6 | 6 | 6 | 6 | 15 | 15 | 15 | 17 | 16 | 20 | 15 | 15 | 15 | 15 | 15 | 16 | 66 | 45 | 49 | 40090 | 45 | 79 | 51 | 46 | 50 | 46 | 47 | 61 |
| 3 | 7 | 7 | 7 | 7 | 7 | 7 | 7 | 7 | 7 | 7 | 8 | 7 | 18 | 15 | 15 | 33 | 17 | 15 | 16 | 15 | 25 | 19 | 15 | 15 | 55 | 48 | 47 | 51720 | 50 | 50 | 46 | 51 | 46556 | 41326 | 57 | 47 |
| 4 | 7 | 7 | 7 | 7 | 7 | 7 | 7 | 7 | 7 | 7 | 7 | 7 | 15 | 15 | 18 | 15 | 15 | 17 | 18 | 15 | 15 | 15 | 16 | 20 | 120 | 59 | 70 | 49 | 50 | 89 | 60 | 57 | 81 | 53 | 46 | 54 |
| 5 | 7 | 7 | 7 | 7 | 7 | 7 | 7 | 7 | 7 | 7 | 7 | 7 | 17 | 15 | 15 | 15 | 15 | 16 | 15 | 15 | 16 | 15 | 16 | 15 | 44 | 45 | 59 | 63 | 45 | 56 | 70 | 55 | 48 | 47 | 49 | 59 |
| 6 | 6 | 6 | 6 | 6 | 6 | 6 | 6 | 6 | 6 | 6 | 6 | 6 | 14 | 14 | 14 | 17 | 27 | 14 | 14 | 14 | 14 | 14 | 15 | 18 | 22509 | 56 | 49 | 45 | 127 | 49 | 37225 | 107 | 59 | 51 | 56 | 51 |
| 7 | 6 | 6 | 6 | 6 | 6 | 6 | 6 | 6 | 6 | 6 | 6 | 6 | 14 | 14 | 15 | 17 | 14 | 14 | 15 | 15 | 15 | 14 | 14 | 14 | 48 | 54 | 48 | 47 | 93 | 45 | 51 | 88 | 56 | 51 | 52 | 48 |
| 8 | 6 | 6 | 7 | 6 | 6 | 6 | 6 | 6 | 6 | 6 | 7 | 6 | 15 | 14 | 15 | 15 | 15 | 15 | 15 | 16 | 14 | 14 | 17 | 15 | 56 | 155 | 46 | 48 | 54 | 44 | 59 | 75 | 43 | 46 | 49 | 43 |
| 10 | 7 | 7 | 7 | 7 | 7 | 7 | 7 | 7 | 7 | 7 | 7 | 7 | 16 | 15 | 18 | 16 | 15 | 15 | 15 | 15 | 16 | 15 | 16 | 15 | 51 | 79 | 90 | 60 | 48 | 65 | 46 | 48 | 55 | 53 | 52 | 77 |
| 11 | 7 | 7 | 7 | 7 | 7 | 7 | 7 | 7 | 7 | 7 | 7 | 7 | 15 | 15 | 15 | 16 | 15 | 24 | 16 | 15 | 15 | 15 | 15 | 15 | 50 | 46 | 65 | 83 | 46 | 115 | 77 | 58 | 46 | 67 | 48 | 54 |
| 12 | 6 | 6 | 6 | 6 | 6 | 6 | 6 | 6 | 6 | 6 | 6 | 6 | 15 | 14 | 21 | 14 | 14 | 14 | 15 | 14 | 14 | 15 | 15 | 14 | 43 | 42 | 55 | 45 | 44 | 62 | 42 | 49 | 148 | 48 | 51 | 52 |
| 13 | 6 | 6 | ## | 6 | 6 | 6 | 6 | 6 | 6 | 6 | 6 | 6 | 15 | 15 | 15 | 15 | 15 | 21 | 15 | 15 | 16 | 15 | 15 | 18 | 81 | 51 | - | 54 | 59 | 78 | 65 | 52 | 53 | 91 | 65 | 57 |
| 14 | 7 | 7 | 7 | 7 | 7 | 7 | 7 | 7 | 7 | 7 | 7 | 7 | 15 | 14 | 14 | 14 | 19 | 15 | 14 | 14 | 14 | 14 | 15 | 14 | 45 | 51 | 47 | 97 | 44 | 50 | 48 | 46 | 44 | 49 | 47 | 49 |
| 15 | 6 | 6 | 6 | 6 | 6 | 6 | 6 | 6 | 6 | 6 | 6 | 6 | 14 | 15 | 14 | 14 | 14 | 14 | 14 | 15 | 15 | 14 | 14 | 22 | 46 | 89 | 44 | 62 | 59 | 62 | 46 | 49 | 50 | 51 | 45 | 50 |

**Table S2 - Cluster extent thresholds for 1st level, native space data of the second fMRI run.** Conditions in which there were no significant clusters are marked with ‘-’.

**0 mm smoothing**

| Contrast | Activation_Regions | Peak_MNI [x, y z] | Peak_T | Cluster_Size |
|---|---|---|---|---|
| looking1_rest-1 | *Fusiform_R (15)* | [26.0, -74.0, -4.0] | 5.89 | 22 |
| naming1_looking-1 | *Cerebellum_6_R (24)* | [20.0, -62.0, -22.0] | 6.14 | 26 |
| naming1_rest-1 | *Postcentral_L (75)* | [-50.0, -12.0, 42.0] | 6.98 | 94 |
| naming1_silent-1 | Precentral_R (34) | [52.0, -6.0, 32.0] | 6.63 | 56 |
| | *Postcentral_R (20)* | | | |
| silent1_rest-1 | *Supp_Motor_Area_L (113)* | [-2.0, 2.0, 62.0] | 8.18 | 153 |

**3 mm smoothing**

| Contrast | Activation_Regions | Peak_MNI [x, y z] | Peak_T | Cluster_Size |
|---|---|---|---|---|
| looking1_rest-1 | Fusiform_R (201) | [32.0, -68.0, -20.0] | 6.42 | 351 |
| | *Cerebellum_6_R (2)* | | | |
| naming1_looking-1 | *Cerebellum_6_L (141)* | [-12.0, -60.0, -24.0] | 6.13 | 155 |
| naming1_rest-1 | Occipital_Mid_L (336) | [-2.0, -88.0, -10.0] | 8.76 | 1160 |
| | *Calcarine_L (94)* | | | |
| naming1_silent-1 | Postcentral_L (285) | [-58.0, -4.0, 30.0] | 8.3 | 386 |
| | *Precentral_L (90)* | | | |
| silent1_rest-1 | Occipital_Mid_L (642) | [-2.0, -92.0, -8.0] | 11 | 4386 |
| | *Calcarine_L (237)* | | | |

**6 mm smoothing**

| Contrast | Activation_Regions | Peak_MNI [x, y z] | Peak_T | Cluster_Size |
|---|---|---|---|---|
| looking1_rest-1 | Fusiform_R (584) | [32.0, -66.0, -20.0] | 6.72 | 1289 |
| | *Cerebellum_6_R (190)* | | | |
| naming1_looking-1 | *Cerebellum_6_L (258)* | [-14.0, -60.0, -22.0] | 5.64 | 294 |
| naming1_rest-1 | *Fusiform_R (922)* | [38.0, -60.0, -2.0] | 9.08 | 7437 |
| naming1_silent-1 | Postcentral_L (712) | [-58.0, -4.0, 32.0] | 9.14 | 1033 |
| | *Precentral_L (228)* | | | |
| silent1_rest-1 | Occipital_Mid_L (1427) | [22.0, -92.0, 6.0] | 12.21 | 10074 |
| | *Occipital_Sup_R (165)* | | | |

**Table S3 - fMRI activation regions.** The table presents the significantly activated regions identified in the second-level analyses. Only the largest clusters are included, with a specific focus on areas exhibiting the highest number of significantly activated voxels and peak voxel locations. Region extraction was performed using the AAL3 atlas. Regions containing the peak voxel of the listed clusters are italicized, while the number of significantly activated voxels is indicated in parentheses in the 'Activated_Regions' column.

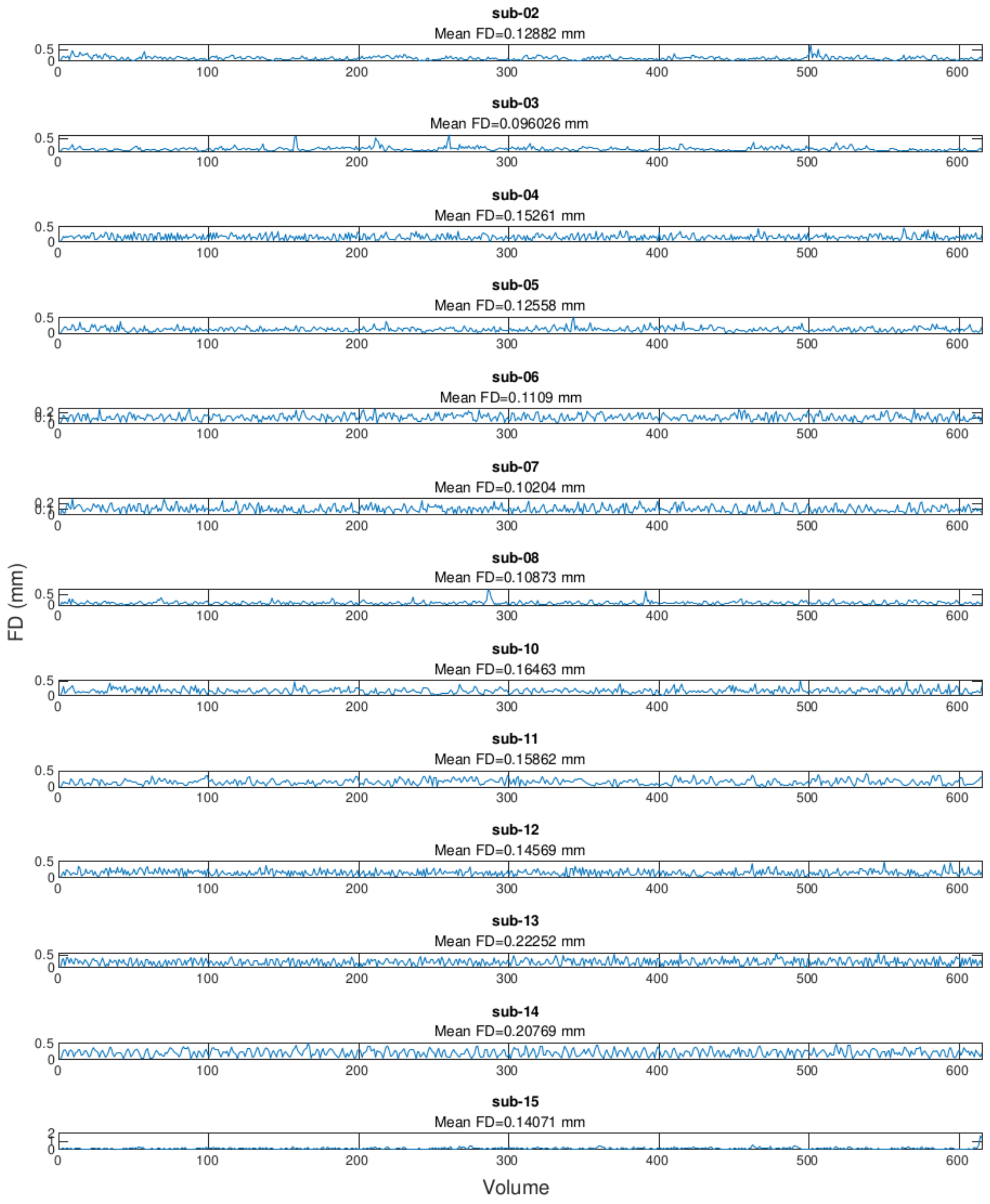


**Fig S1 - Framewise displacement of fMRI data (first run).**

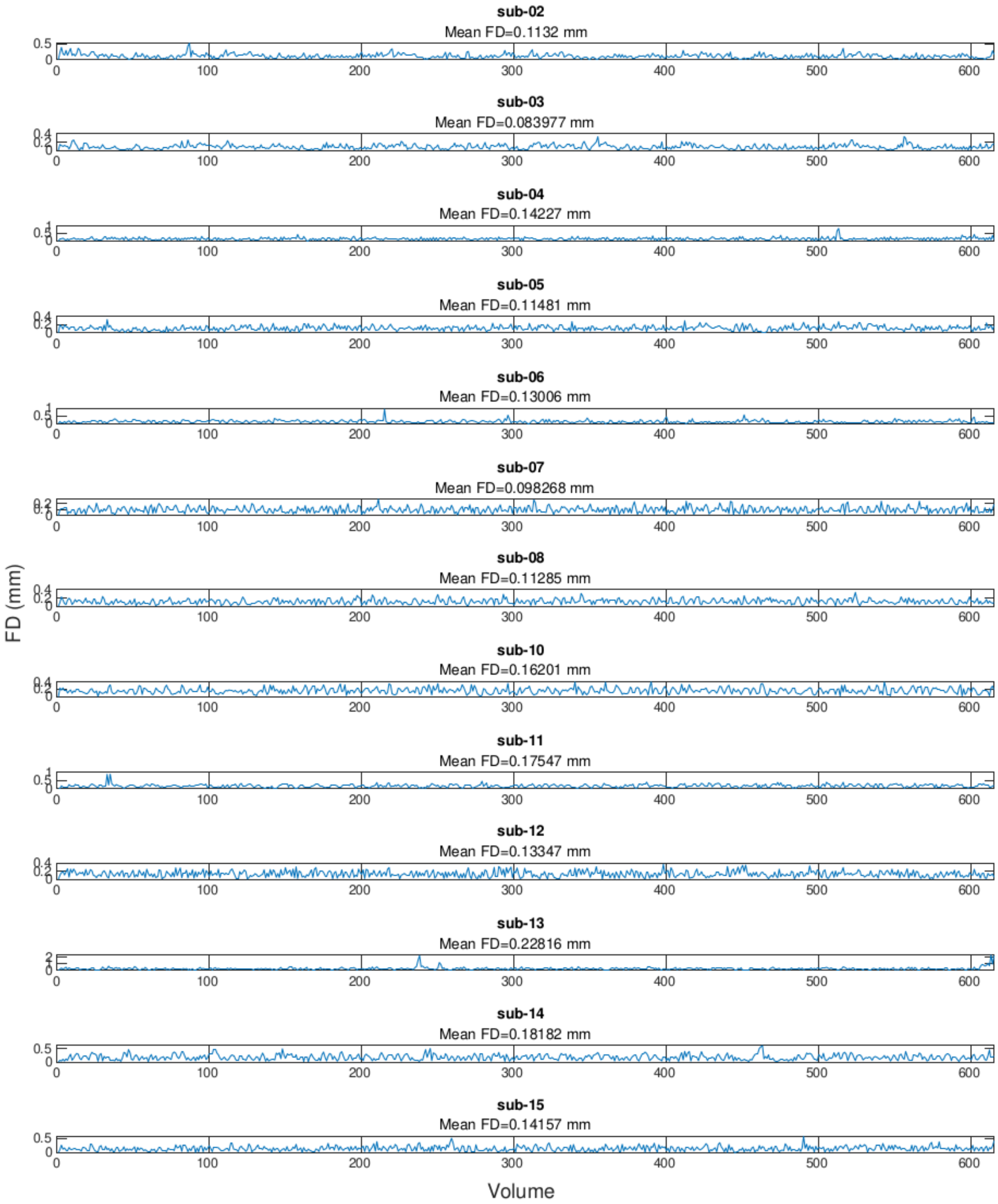


**Fig S2 - Framewise displacement of fMRI data (second run).**

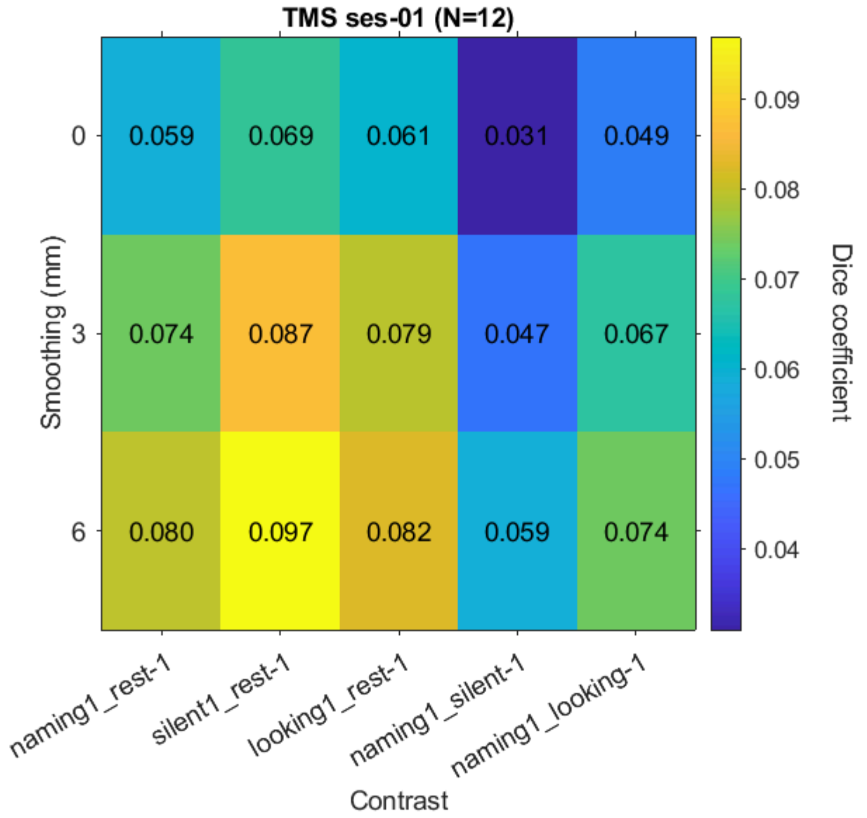


**Fig S3 - Dice coefficients.** The figure represents Dice coefficients averaged across subjects (N=12), in each smoothing level and contrast.

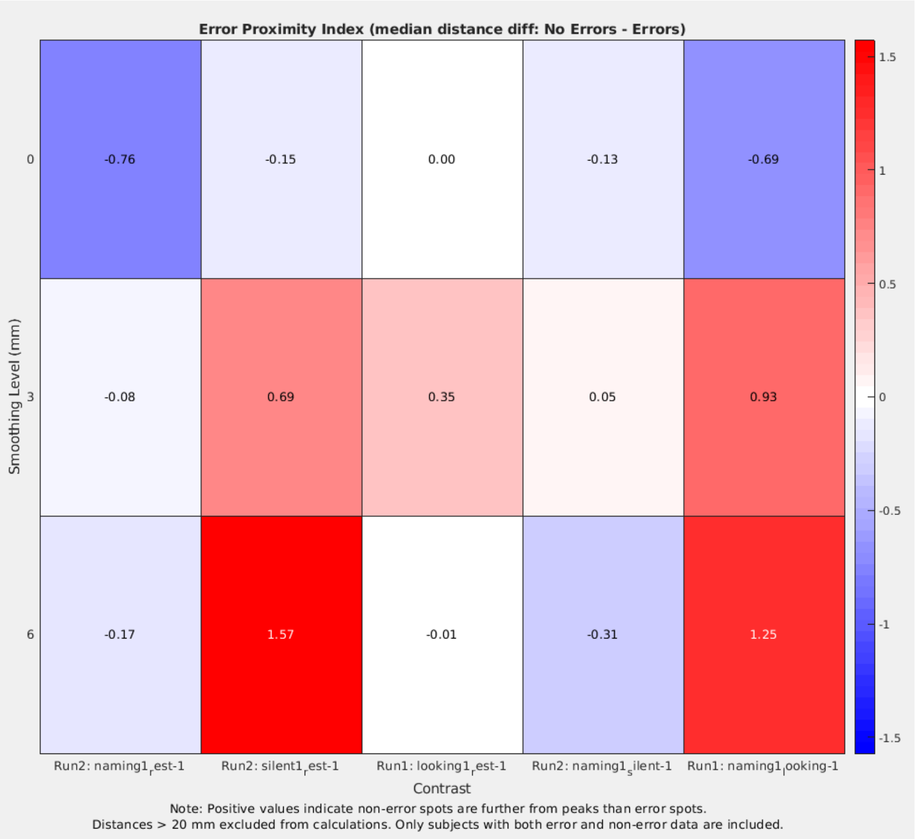


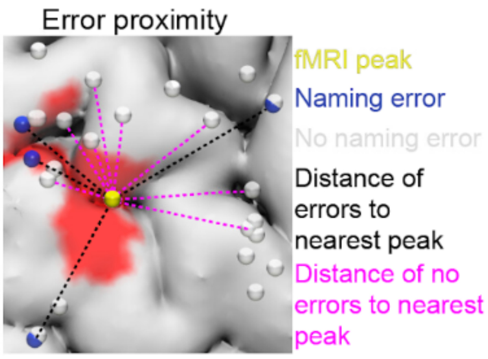


**Fig S4 - Naming Error Proximity Indices (Euclidean distances)**. We extracted the peak coordinates of significant first-level clusters for each contrast and smoothing level. We then searched for the closest cluster peak relative to each TMS coordinate. To measure spatial proximity of TMS and fMRI mapping, we calculated the Euclidean distances between error-producing TMS sites and those that did not produce naming errors. Lastly, we computed the *Naming Error Proximity Index* as the difference between the median distance of non-error-producing sites to the closest fMRI peak and the median distance of error-producing sites to the closest peak, using the formula:

$$\text{Naming Error Proximity Index} = \\ \text{Median}(\text{noErrorSites vs closestPeak}) - \\ \text{Median}(\text{errorProducingSites vs closestPeak})$$

Thus, a higher Naming Error Proximity Index value indicates a better correspondence between TMS-induced naming errors and fMRI data.
